# Safety first: should the high tolerability of intramuscular anti-spike COVID-19 monoclonal antibody change our expectations of vaccine safety?

**DOI:** 10.64898/2026.05.08.26352596

**Authors:** David Putrino, Allison Curtis, Meredith Leston, Ilker Yalcin, Rachael Gerlach, Marc Elia, Michael Mina

## Abstract

**Introduction:** Public and regulatory scrutiny of immunization safety has intensified in recent years. The COVID-19 pandemic has been instrumental in this. The accelerated timeline of COVID-19 vaccine development combined with the amplification of resultant side effects have proven corrosive to confidence. Unsurprisingly, COVID-19 vaccine uptake has declined year-on-year. This conflicts with the threat that infection still presents: predictors and prognoses of post-acute complications remain uncertain. Restoring public trust in these technologies will require meaningful progress in the availability and accessibility of clinical safety and pharmacovigilance data.

**Methods:** Expanding upon recent comparisons of COVID-19 vaccine reactogenicity, we present a post-hoc safety analysis of adintrevimab, an intramuscular (IM) anti-SARS-CoV-2 spike recombinant investigational monoclonal antibody (mAb) for the pre-exposure and post-exposure prophylaxis of COVID-19, as assessed by the multi-center, double-blind, Phase 2/3 randomized placebo-controlled EVADE study (NCT04859517). Exploratory endpoints included the incidence of ≥1 systemic symptoms within 7 days of study drug administration as well as symptom number, duration and severity. Safety reporting encompassed solicited and unsolicited treatment-emergent adverse events (TEAEs), serious adverse events (SAEs), vital signs, and clinical laboratory assessments.

**Results:** EVADE study participants (n=2582) were randomized between April 2021 – January 2022. Baseline characteristics were balanced across treatment groups. Within the 7 day post-dose period, 25/1241 (2.0%) of adintrevimab recipients and 12/1242 (1.0%) of placebo recipients reported at least one systematic TEAE. Multiple systemic TEAEs were less prevalent, with 0.3% and 0.1% reporting two systemic TEAEs, and 0.1% and 0.1% reporting three TEAEs in adintrevimab and placebo groups, respectively. The majority of TEAEs reported were mild to moderate in severity, primarily involving headache (0.4% adintrevimab, 0.8% placebo), fatigue (adintrevimab 0.4%, placebo 0.2%), and nausea/vomiting (adintrevimab 0.4%, placebo 0.1%). For those participants who experienced any TEAEs in the 7 day post-dose period, mean (+/-standard deviation) number of systemic symptoms was 1.2 (0.5) for adintrevimab and 1.3 (0.6) for placebo with symptoms consistently resolving within 3 days.

**Conclusions:** Increased expectations for pharmaceutical safety data generation are to be welcomed, offering patients the information they need to appropriately weigh the benefits and risks of any novel therapeutic. These analysis results support the high tolerability of IM-administered adintrevimab, with reactogenicity data broadly comparable to placebo. While the co-administration of vaccines and monoclonal antibodies limit direct comparisons between historical safety reports, findings such as these demonstrate the potential clinical value of controlled head-to-head studies such as the anticipated LIBERTY trial.

## INTRODUCTION

Since May of 2023, the widespread availability of COVID-19 immunizations, therapeutics and their combined impacts on acute infection outcomes have been cited to justify World Health Organization (WHO) downgrade of COVID-19 from a Public Health Emergency of International Concern [1]. Indeed, acute COVID-19 infection now rarely results in hospitalization, and cases primarily present as mild or self-limiting [2].

However, the burden of COVID-19 illness remains significant [3], and is accompanied by mounting evidence of post-acute complications, including protracted sequelae [4], new-onset etiologies [5-7], and exacerbations of pre-existing conditions [8-10], which are not yet completely understood and communicated [11]. Emerging associations between SARS-CoV-2 infection’s hyperinflammatory properties [12] and downstream autoimmune [13], oncogenic [14-16], and neurodegenerative consequences [17-19] consequences continue to be incompletely characterized.

Vaccination against SARS-CoV-2 has been one of the mainstays for prevention of severe COVID-19. However, the past years have demonstrated increasing public concern regarding the safety and tolerability risks of COVID-19 vaccination [20, 21] with expected patterns of vaccine refusal that, unlike influenza, appear invariant to increased threat of infection [22, 23]. Increasingly, public awareness of side effects, which can drive systemic effects such as fever and soreness, and can drive days missed from work or school, is contributing to lower uptake [24]. Vaccine refusal also extends to populations with well-established vulnerability to COVID-19 infection where confirmatory safety data are perceived as lacking [25, 26], and uptake now hovers between 20-30% and has been trending downwards year on year [27].

The WHO’s ‘3Cs framework’ defines the influence of public *confidence, complacency*, and *convenience* on vaccine uptake [28]. Evaluation through this framework suggests that the current socio-medical landscape is highly antagonistic to COVID-19 prophylaxis. Concordantly, US regulatory agencies have recently introduced more stringent reactogenicity reporting requirements for immunologic products seeking licensure [29, 30].

The COMPARE study [31, 32] is a head-to-head Phase 4 study that delineates the relative reactogenicity of contemporary mRNA-based versus protein-based approaches to COVID-19 vaccination in terms of Grade 2/3 systemic adverse events, among other analyses, in the 7 days post vaccine boost. Results of COMPARE indicated very high rates of systemic adverse reactions to the vaccine, including 84% and 92% of participants reporting at least one systemic adverse reaction within 7 days post inoculation with the protein-based and mRNA-based vaccines, respectively, lasting an average of 3.1 and 3.5 days. Despite these high rates, these results were taken to conclude a superior tolerability profile of the protein-based vaccine.

Similar comparative tolerability studies of monoclonal antibody-based approaches to COVID-19 prophylaxis have yet to be performed or positioned head-to-head against vaccine options. While safety data are reported at each phase of clinical development, these agents have not been incorporated into large-scale pharmacovigilance platforms including Vaccine Adverse Event Reporting System (VAERS) or V-Safe, or postmarketing efforts [33, 34]. This, combined with aforementioned changes to regulators’ safety expectations, and the WHO’s 3Cs framework, places unique onus on industry to more proactively assess and publicize the tolerability of these therapies. Rigorous and continuous monitoring, as well as data that extend beyond cross-trial comparison, are essential for empowering shared clinical decision-making.

As such, this work presents a post-hoc evaluation of the rates of systemic side effects associated with adintrevimab, an anti-SARS-CoV-2 spike recombinant investigational monoclonal antibody [35] as assessed in the EVADE study (NCT04859517), a multi-center, double-blind, Phase 2/3 randomized, placebo-controlled trial of the IM administration of adintrevimab. While the primary analyses of EVADE focused on the prevention of symptomatic COVID-19 and overall safety, the frequency and timing of systemic adverse events immediately following mAb administration warrant further evaluation. The EVADE dataset was selected due to adintrevimab’s IM formulation, which is consistent with its successor’s route of administration, the SARS-CoV-2 directed investigational monoclonal antibody, VYD2311. As well as providing an important precedent for monoclonal antibody benefit-risk profiling, these tolerability data will provide a benchmark for the ongoing clinical trials of VYD2311, including DECLARATION (NCT07298434), an ongoing Phase 3 randomized, triple-blind, placebo-controlled clinical trial to evaluate the efficacy and safety of VYD2311 for the prevention of COVID-19.

## METHODS

### Study Design

This post hoc analysis was conducted using post-dose tolerability data from EVADE, a multi-center, double-blind, placebo-controlled, randomized Phase 2/3 study designed to evaluate the safety and efficacy of adintrevimab for pre-exposure and post-exposure prophylaxis of COVID-19. Full details of the EVADE study design, eligibility criteria, and primary analysis have been reported previously [35].

### Patient Consent

Written informed consent was obtained from all participants. The study was conducted in accordance with the International Conference on Harmonisation Good Clinical Practice guidelines, the Declaration of Helsinki, and all applicable regulations. The protocol was reviewed and approved by an institutional review board or ethics committee in each country. In the United States, Advarra (Columbia, MD, USA) approved the study for all sites.

### Analysis Population

The safety analysis population for this post hoc assessment included all randomized participants who received at least one dose of study treatment (safety population). Unsolicited treatment-emergent adverse events (TEAEs) occurring within the predefined post-dose window (7 days) were included. Solicited injection-site reactions (ISRs) and hypersensitivity reactions were recorded through Day 4.

### Post Hoc Analysis Objectives

The objective of this post hoc analysis was primarily to characterize the incidence and timing of systemic adverse events occurring within the first week following administration of study mAb treatment. Additional exploratory analyses included: 1) Incidence of experiencing at least one systemic symptom within 7 days of study drug dose, 2) Symptom number, duration and severity. The post-dose risk window was defined as Day 0 (day of dosing) through Day 7.

In all participants who received study drug, safety was assessed, including TEAEs, serious adverse events (SAEs), vital signs, and clinical laboratory assessments. TEAEs in the original study was defined as any AE that had an onset during or after the administration of study drug through to Month 14, or any preexisting condition that has worsened during or after the administration of study drug through Month 14. TEAEs were captured at in-person study visits throughout the study periods. However, for this post-hoc analysis, any unsolicited systemic TEAEs that occurred within the predefined post-dose window (7 days) were included. These data were captured by study sites on Day 8 during the in-person study visit. The systemic adverse events were defined as TEAEs reflecting generalized or non–injection-site responses, including constitutional symptoms (e.g., fever, chills, fatigue, headache, myalgia), and gastrointestinal symptoms (diarrhea, nausea, vomiting).

Solicited injection-site reactions (ISRs; injection site pain, tenderness, erythema/redness, and/or swelling) and hypersensitivity reactions were recorded through Day 4.

All adverse events were coded using the Medical Dictionary for Regulatory Activities (MedDRA), version 24, and categorized by system organ class and preferred term.

### Severity Assessment

The intensity of all unsolicited TEAEs was graded by the investigator according to the DAIDS Table for Grading the Severity of Adult and Pediatric Adverse Events [36]. The DAIDS grading table provides a TEAE severity grading scale ranging from grades 1 to 5. When changes in the intensity of a TEAE occurred more frequently than once a day, the maximum intensity was noted. If the intensity category changed over a number of days, those changes were recorded separately (with distinct onset dates). ISRs (solicited AEs) were considered related to study drug. The investigator assessed causality for all unsolicited TEAEs.

### Statistical Analyses

All post hoc analyses were analyzed using descriptive statistics. Unsolicited systemic adverse events and solicited ISRs were summarized by treatment group using counts and percentages within the 7 day post-dose window. Participants experiencing multiple events were counted daily per preferred term. No formal hypothesis testing was performed, consistent with the exploratory nature of this post hoc analysis. Statistical analyses were performed using SAS Version 9.4.

## RESULTS

### Participants

EVADE study participants were randomized between April 27, 2021 and January 11, 2022. The post hoc safety analysis set included 1241 participants in the adintrevimab group and 1242 in the placebo group. As reported previously, baseline characteristics were balanced across treatment groups [35].

### Reactogenicity Assessment

Reactogenicity within 7 days post study drug (adintrevimab vs. placebo) administration was evaluated to characterize early tolerability. During the 7 day post-dose period, at least one systemic TEAE was reported in 25/1241 (2.0%) of participants who received adintrevimab and 12/1242 (1.0%) of participants in the placebo group (Table 1). A smaller proportion experienced multiple systemic TEAEs, with 0.3% and 0.1% reporting two systemic TEAEs, and 0.1% and 0.1% reporting three in adintrevimab and placebo groups, respectively. Across both adintrevimab and placebo groups, most systemic TEAEs were mild to moderate (Grade 1 or 2) in severity (Table 2).

**Table 1:** Incidence of Systemic and Local Treatment Emergent Adverse Events Within 7 days post-administration.

| Category | Adintrevimab<br>(N=1241) | Placebo<br>(N=1242) |
| --- | --- | --- |
| Systemic TEAEs, n (%) |  |  |
| 0 | 1216 (98.0) | 1230 (99.0) |
| 1 | 21 (2.0) | 10 (0.8) |
| 2 | 3 (0.3) | 1 (0.1) |
| 3 | 1 (0.1) | 1 (0.1) |
| 4 | 0 | 0 |
| 5 | 0 | 0 |
| 6 | 0 | 0 |
| 7 | 0 | 0 |
| Injection Site Reactions, n (%) |  |  |
| Grade 1 | 83 (6.7) | 74 (6.0) |
| Grade 2 | 12 (1.0) | 15 (1.2) |
| Grade 3 | 1 (0.1) | 3 (0.2) |
| Grade 4 | 0 | 0 |

**Table 2:**
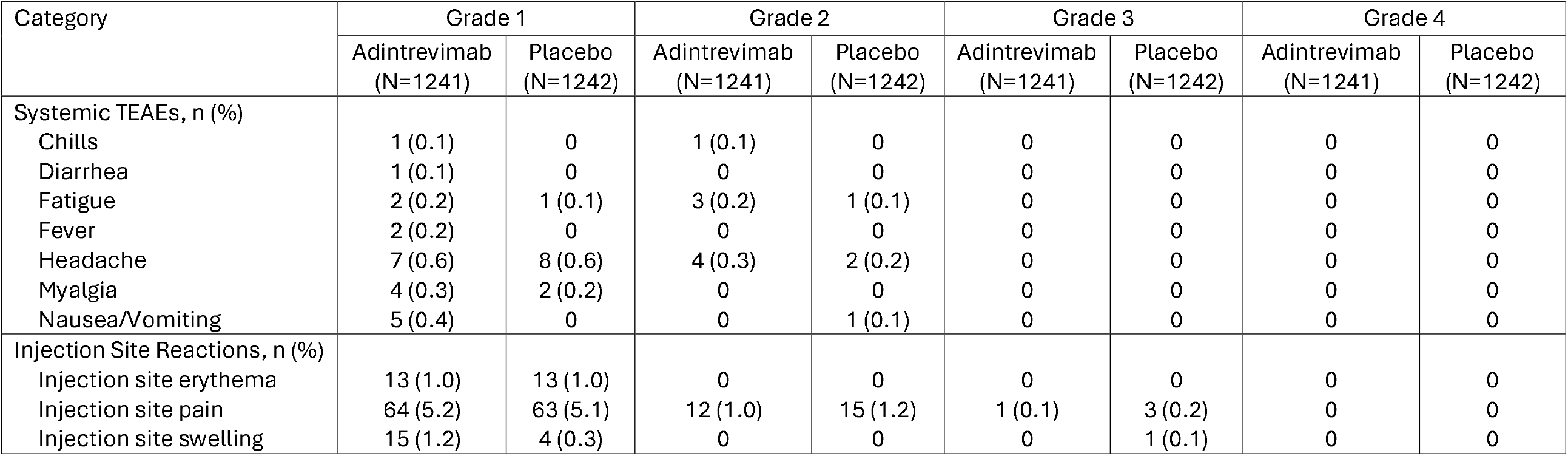
Summary of Systemic and Local Treatment-Emergent Adverse Events by Severity.

| Category | Grade 1 |  | Grade 2 |  | Grade 3 |  | Grade 4 |  |
| --- | --- | --- | --- | --- | --- | --- | --- | --- |
|  | Adintrevimab<br>(N=1241) | Placebo<br>(N=1242) | Adintrevimab<br>(N=1241) | Placebo<br>(N=1242) | Adintrevimab<br>(N=1241) | Placebo<br>(N=1242) | Adintrevimab<br>(N=1241) | Placebo<br>(N=1242) |
| Systemic TEAEs, n (%) |  |  |  |  |  |  |  |  |
| Chills | 1 (0.1) | 0 | 1 (0.1) | 0 | 0 | 0 | 0 | 0 |
| Diarrhea | 1 (0.1) | 0 | 0 | 0 | 0 | 0 | 0 | 0 |
| Fatigue | 2 (0.2) | 1 (0.1) | 3 (0.2) | 1 (0.1) | 0 | 0 | 0 | 0 |
| Fever | 2 (0.2) | 0 | 0 | 0 | 0 | 0 | 0 | 0 |
| Headache | 7 (0.6) | 8 (0.6) | 4 (0.3) | 2 (0.2) | 0 | 0 | 0 | 0 |
| Myalgia | 4 (0.3) | 2 (0.2) | 0 | 0 | 0 | 0 | 0 | 0 |
| Nausea/Vomiting | 5 (0.4) | 0 | 0 | 1 (0.1) | 0 | 0 | 0 | 0 |
| Injection Site Reactions, n (%) |  |  |  |  |  |  |  |  |
| Injection site erythema | 13 (1.0) | 13 (1.0) | 0 | 0 | 0 | 0 | 0 | 0 |
| Injection site pain | 64 (5.2) | 63 (5.1) | 12 (1.0) | 15 (1.2) | 1 (0.1) | 3 (0.2) | 0 | 0 |
| Injection site swelling | 15 (1.2) | 4 (0.3) | 0 | 0 | 0 | 1 (0.1) | 0 | 0 |

The most frequently reported systemic TEAEs were headache (adintrevimab 0.9%, placebo 0.8%), fatigue (adintrevimab 0.4%, placebo 0.2%), and nausea/vomiting (adintrevimab 0.4%, placebo 0.1%) (Table 2).

For those participants who experienced any TEAE within 7 days post-dose, mean (+/-standard deviation) number of systemic symptoms was 1.2 (0.5) for adintrevimab and 1.3 (0.6) for placebo. Most resolved within 3 days for both groups (Table 3).

**Table 3:** Summary of Systemic and Local Treatment-Emergent Adverse Events by Duration.

| Category | 1 Day |  | 2 Days |  | 3 Days |  | ≥ 4 Days |  |
| --- | --- | --- | --- | --- | --- | --- | --- | --- |
|  | Adintrevimab<br>(N=1241) | Placebo<br>(N=1242) | Adintrevimab<br>(N=1241) | Placebo<br>(N=1242) | Adintrevimab<br>(N=1241) | Placebo<br>(N=1242) | Adintrevimab<br>(N=1241) | Placebo<br>(N=1242) |
| Systemic TEAEs, n (%) |  |  |  |  |  |  |  |  |
| Chills | 1 (0.1) | 0 | 1 (0.1) | 0 | 0 | 0 | 0 | 0 |
| Diarrhea | 1 (0.1) | 0 | 0 | 0 | 0 | 0 | 0 | 0 |
| Fatigue | 2 (0.2) | 0 | 1 (0.1) | 0 | 0 | 0 | 1 (0.1) | 2 (0.2) |
| Fever | 2 (0.2) | 0 | 0 | 0 | 0 | 0 | 0 | 0 |
| Headache | 2 (0.2) | 4 (0.3) | 3 (0.2) | 2 (0.2) | 1 (0.1) | 0 | 5 (0.4) | 3 (0.2) |
| Myalgia | 3 (0.2) | 0 | 0 | 0 | 0 | 0 | 0 | 2 (0.2) |
| Nausea/Vomiting | 4 (0.3) | 0 | 0 | 0 | 1 (0.1) | 0 | 0 | 1 (0.1) |
| Injection Site Reactions, n (%) |  |  |  |  |  |  |  |  |
| Injection site erythema | 9 (0.7) | 8 (0.6) | 3 | 4 (0.3) | 0 | 1 (0.1) | 1 (0.4) | 1 (0.1) |
| Injection site pain | 57 (4.6) | 61 (4.9) | 13 (1.0) | 10 (0.8) | 4 (0.3) | 7 (0.6) | 4 (0.3) | 5 (0.4) |
| Injection site swelling | 10 (0.8) | 6 (0.5) | 2 (0.2) | 0 | 1 (0.1) | 0 | 2 (0.2) | 0 |

Overall, solicited ISRs Grade 1 or higher were reported in 7.5% of participants in the adintrevimab group and 7.3% for placebo (Table 1). A smaller proportion of ISRs were Grade 2 (1.0% adintrevimab, 1.2% placebo), and Grade 3 (0.1% adintrevimab, 0.2% placebo). Most ISRs were resolved within 3 days (Table 3).

## DISCUSSION

Transparent benefit-risk profiling is foundational to medical ethics and patient trust: uncertainty risks can produce irreparable damage to both. Increasing public demand for safety and tolerability reporting is therefore to be welcomed, especially in immunization deployed for the protection of well subjects. In this post-hoc analysis of EVADE safety data, systemic TEAEs and ISRs occurring within the first week of a single IM dose of the SARS-CoV-2 directed mAb adintrevimab were evaluated to further characterize early tolerability. Overall, immunization through this means was extremely well tolerated, with low incidence of local ISRs (7.5%) and systemic reactions (2.0%), and lower yet when adjusted for placebo control. These results were only marginally higher than the placebo control, highlighting the minimal reactogenicity of this approach to COVID-19 prophylaxis.

The favorable profile demonstrated here is not unique to these data, however, but appears characteristic of passive immunization and its avoidance of immune activation pathways [37]. Safety data embedded within efficacy evaluations of other historical low dose SARS-CoV-2 directed mAbs sotrovimab, bebtelovimab, and sipavibart are broadly consistent with the data reported here [38-40]. Unlike vaccination, mAb-induced immunity does not elicit innate systems, antigen presentation, or adaptive immune expansion – all of which are key drivers of reactogenicity and systemic symptoms, which are highly burdensome for a seasonal prophylactic measure [37].

Such comparisons between vaccine and mAb pathways are important hypothesis-generating exercises that should not be avoided due to the sensitivities associated with immunization or protectiveness for one modality over the other [31]. Instead, as an adjunct to our primary analysis, we present an exploratory extrapolative exercise comparing COMPARE and EVADE studies’ systemic tolerability with contemporary COVID-19 symptom duration to estimate total symptomatic days avoided by each modality. We modeled these outputs for a 5% attack rate for unimmunized at four levels of immunization efficacy for a single dose: 25%, 50%, 75%, and 100% of each modality. At the population level, overall symptomatic burden reflects both reactogenicity and COVID-19 illness. We illustrate here that, irrespective of efficacy scenario, reduction in systemic TEAEs would translate to pronounced improvement to overall symptom burden (Figure 1).

**Figure 1:**
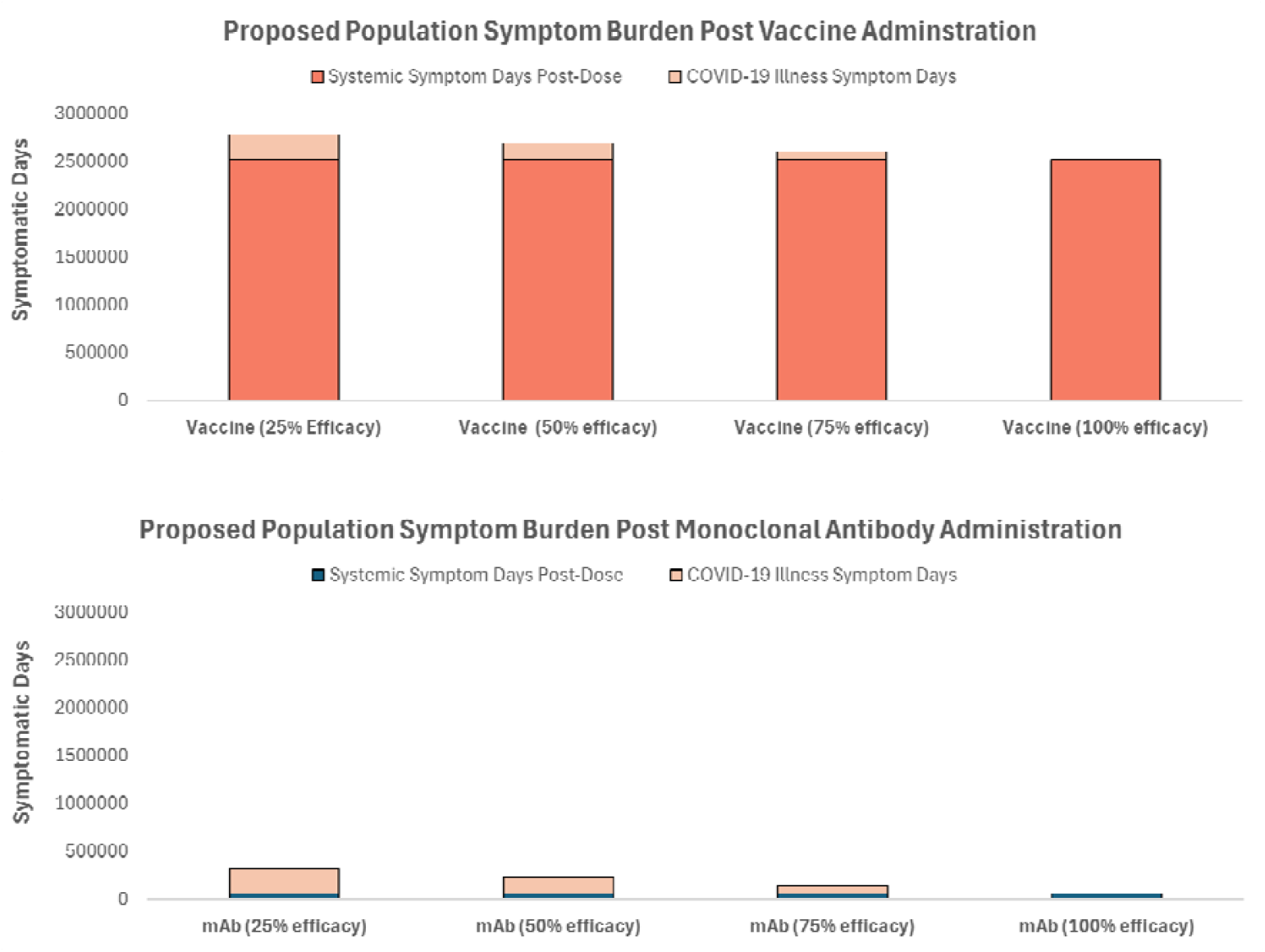
Extrapolated Symptom Burden (Systemic TEAEs + COVID-19 Illness Days) per 1 million individuals. Total Symptomatic Days = N × [(Fraction with systemic TEAEs × Average Treatment-induced Symptom Duration) + ((1-Treatment Efficacy) × COVID-19 Attack Rate × Average Disease Symptom Duration)] Where N = 1 million individualsCOVID-19 Attack Rate = 5% for unimmunized Fraction with systemic TEAEs: 0.84 (vaccine), 0.02 (monoclonal antibody, mAb) Average Treatment-induced symptom duration: 3 days (vaccine), 3 days (mAb) COVID-19 attack rate for unimmunized population: 5% Average Disease Symptom Duration: 7 days

While these hypotheticals are of academic interest, they do not reflect current clinical reality: the co-administration of such technologies and reliance on accurate patient self-report [41] limits the comparability and scientific rigor of these datasets. Head-to-head comparisons between these modalities will remain contentious without highly controlled, blinded designs.

To address this unmet need, the Phase 3 LIBERTY trial will evaluate the comparative and combined safety, tolerability, and pharmacokinetics of VYD2311 versus mRNA COVID-19 vaccination in healthy adults [42]. Thoughtful delineation between study arms will support comparative benefit-risk profiling between these modalities for the first time. Such work is a valuable template for identifying the optimal method for immunization and the first-line potential of monoclonal antibody-based approaches.

Meanwhile, the ongoing Phase 3 DECLARATION trial will establish the safety profile of a multi-dose schedule across a variety of clinical and non-clinical populations [43]. Initial safety findings from the first-in-human evaluation of high dose VYD2311 have been encouraging with reported adverse events predominantly mild to moderate and largely limited to injection-site or infusion-related reactions [44].

## LIMITATIONS

There are several limitations that should be considered when interpreting the findings. The evaluation of systemic adverse events within the early post-dose window was conducted as a post hoc analysis of the EVADE study and therefore not prospectively powered to detect differences in tolerability outcomes. Reliance on unsolicited participant-reported symptoms may introduce reporting bias for adverse events and some may be underestimated. Discussion comparisons with vaccine reactogenicity data are indirect and constrained by differences in study design, populations, and type of symptom reporting.

## CONCLUSIONS

This post-hoc analysis of EVADE safety data adds to mounting evidence for the tolerability of monoclonal antibody approaches to COVID-19 prevention and underscores the need for prospective head-to-head safety profiling of novel immunizations to eliminate the uncertainties of cross-trial comparisons and to establish reliable benchmarks for net symptomatic benefits. Work such as this will be essential for assessing the unique value of a monoclonal antibody-inclusive immunization paradigm.

## Data Availability

All data produced in the present work are contained in the manuscript.

## Acknowledgements

Financial support: This work was supported by Invivyd, Inc.

## Data Sharing Statement

All data produced in the present work are contained in the manuscript.

